# Pulmonary tuberculosis prediction using CAD4TB artificial intelligence (computer-aided detection for tuberculosis) based on thoracic x-ray photos among Indonesian subjects in hospital

**DOI:** 10.1101/2025.04.20.25326134

**Authors:** Erlina Burhan, Maryastuti, Listi Wulandari, Diah Handayani, Salsabila Rezkia Andini, Anandya Naufal Rahadhi, Gde Ngurah Irfan Bhaskara, Khansa Putrirana, Ariestiana Ayu Ananda Latifa, Ahmad Fadhil Ilham, Ihya Akbar, M Prasetio Wardoyo

**Author notes:** Corresponding author: Prof. Erlina Burhan, Department of Pulmonology & Respiratory Medicine, Faculty of Medicine Universitas Indonesia - Persahabatan Central General Hospital, Jalan Persahabatan Raya No. 1, Rawamangun, Pulogadung, East Jakarta 13230, Indonesia.

## Abstract

Tuberculosis remains a major global health concern, particularly in high-burden countries where early detection is essential but often limited by insufficient radiological expertise. This study evaluated the diagnostic performance of a computer-aided detection system, CAD4TB, in interpreting chest X-ray images of suspected tuberculosis cases in a hospital setting in Indonesia. Using a retrospective, cross-sectional design, we analyzed chest radiographs from over 1,100 adult patients drawn from the national tuberculosis database. Images were processed using the CAD4TB system and independently reviewed by two experienced radiologists. Bacteriological test results were used as the diagnostic reference standard. At a CAD4TB index cutoff of 60, the tool achieved a sensitivity of 81.04% and a specificity of 63.80%. In comparison, radiologist interpretations achieved a sensitivity of 88.20% and a specificity of 58.18%. Subgroup analyses revealed improved diagnostic performance in individuals without pleural effusion, with CAD4TB sensitivity rising to 83.65% and radiologist sensitivity to 90.35%. CAD4TB also showed consistent specificity advantages across clinical subgroups, including those with prior tuberculosis history and HIV-negative status. These findings support the potential role of CAD4TB in assisting radiologists within hospital settings, especially in high-burden areas. Its time-efficiency and ease of use make it a valuable tool for integration into tuberculosis triage systems, particularly where patient complexity varies and access to expert readers is limited.

**Author Summary:** Tuberculosis is still one of the world’s leading infectious diseases, especially in countries like Indonesia where many cases go undetected due to limited access to expert medical imaging professionals. In this study, we explored whether a computer system called CAD4TB, which uses artificial intelligence to read chest X-rays, could help identify people with tuberculosis in a hospital setting. We compared its performance to experienced radiologists and used laboratory tests to confirm the results. We found that CAD4TB was able to detect tuberculosis with similar accuracy to human experts in many cases, especially among patients without complications like fluid in the lungs. Although expert radiologists remain slightly more accurate overall, CAD4TB performed well enough to suggest that it could be used to support hospital teams—especially where there are not enough trained readers. Because this system can analyze X-rays quickly and consistently, we believe it could be useful in busy hospitals and in regions with high numbers of tuberculosis cases. Our findings may help health programs consider how digital tools like CAD4TB can be integrated into screening and diagnosis strategies to improve early detection and treatment.

## Introduction

Tuberculosis (TB) remains a significant global health challenge, with approximately one-third of the world’s population estimated to be infected by Mycobacterium tuberculosis. In Indonesia, the TB incidence rate is 842,000 cases annually, with 32% of these cases either undetected or unreported.[1] Consequently, early detection and treatment of TB are global priorities.

Chest x-ray (CXR) role in identifying TB lung pathology was often hampered by intra-reader variability and a shortage of skilled radiologists’ in countries with high TB burdens.[2] Progress in the field of artificial Intelligence (AI) helped solving those challenges. Current studies have utilized AI-powered Computer-aided Detection (CAD), such as CAD4TB, to assist in diagnosing pulmonary TB using CXRs, improving its accessibility, affordability, and accuracy. However, AI has not yet been fully implemented for TB diagnosis in Indonesia.[2] Therefore, using previous CXR imaging and retrospective data of suspected TB patients, this study aims to evaluate CAD4TB software in detecting tuberculosis compared to the radiologists’ interpretation of CXR images, with bacteriological assay of TB as the reference standard.

## Materials and Methods

### A. Study Design and Participants

This cross-sectional study was conducted at Persahabatan Hospital using the Tuberculosis Information System (*Sistem Informasi Tuberkulosis,* SITB) database. The inclusion criteria for this study are suspected TB patients, which are the persons with either one of four classic TB symptoms (productive cough, fever, weight loss, night sweats) or other reason to be suspected with TB, assessed between 2018-2024 and aged ≥ 18 years. Specifically, for the TB-positive group, bacteriological sample collection should be taken within 7 days before or after CXR, and the bacteriological assay (at least one of acid-fast bacilli (AFB) smear, molecular rapid diagnostic (MRD), and culture) must be found positive. Patients were excluded if they are pregnant, lack bacteriological assay results in their medical records, or have CXR results that do not meet the data interpretation. Subjects’ inclusion was done using total sampling approach, with an estimated total of 1,100 samples, comprising 500 TB-positive subjects and 600 randomly selected TB-negative subjects.

### B. CXRs readings

All CXR data from each sample was anonymized using specific patient IDs. The data was analyzed using CAD4TB version 7 in collaboration with Delft Imaging System Netherlands to generate diagnostic predictions in the form of an index. Based on the best performing cut-off reported on CAD4TB White Paper, an index ≥60 was classified as TB Positive; otherwise, the patient was classified as TB Negative.[3,4] Subsequently, two radiologists’ with clinical experiences of more than five years will independently read the x-ray results and interpret them according to the ’Tanzanian X-ray score’ by Breuninger et al,[5] that is 1) normal (scored 0), 2) abnormal, not suggestive of active TB (scored 1), 3) abnormal, consistent with active TB but TB sequelae or other lung pathology possible (scored 2), and 4) abnormal, highly suggestive of active TB (scored 4). Radiologists’ scoring of 0 and 1 was classified as TB Negative, while the scores of 2 and 3 was classified as TB Positive. Both of these assessment results were then to be confirmed with bacteriological assay.

### C. Data analysis

The confidentiality of the CAD4TB data was maintained by storing the materials on a password-protected device. Firstly, clinical characteristics of subjects was summarized, including symptoms, HIV status, prior TB history, and physical examinations such as weight, height, and BMI. For diagnostic analysis, a microbiological confirmation of Mycobacterium tuberculosis was used as the reference standard to evaluate the diagnostic accuracy of these methods in diagnosing pulmonary tuberculosis.

Statistical analysis was done using R-4.3.1. The receiver operating characteristic (ROC) curve was produced, displaying the area under the curve (AUC) of CAD4TB and radiologists’ interpretation score using pROC package.[6] The AUC of both CAD4TB and radiologists’ interpretation were compared using DeLong test. Diagnostic accuracy parameters of CAD4TB and radiologists’ interpretation, consisted of sensitivity, specificity, positive predictive value (PPV), negative predictive value (NPV), positive likelihood ratio (LR), and negative LR were calculated along with 95% Confidence Intervals (CI). Then, McNemar’s test was used for comparing the sensitivity and specificity of CAD4TB to radiologists’ interpretation, along with relative predictive values approach for comparing PPV and NPV and DLR regression model for comparing positive and negative LR. These comparisons were conducted using DTComPair package.[7–9] From the comparison of CAD4TB with radiologists’ interpretation, two other cutoffs were determined based on where sensitivity or specificity of CAD4TB aligns with radiologists’ interpretation. Subsequently, the analysis of diagnostic parameters at that cutoff point was conducted. Furthermore, subgroup analysis was carried out to determine how factors such as pleural effusion, HIV, and previous TB history influence the accuracy of CAD4TB diagnostic parameters.[10]

### D. Ethical considerations

The RAIT Study was approved by the Ethical Committee Review Board of the Persahabatan Hospital, Indonesia, with the Ethical Clearence No. 115/KEPK-RSUPP/07/2023.

## Results

### A. Subjects’ Characteristics

A total of 1122 retrospective subjects, consisted of 605 TB negative subjects and 517 TB positive subjects, were retrospectively enrolled in the study. Subject screening process, including numbers and reasons of exclusion, was described in Figure 1.

**Figure 1.**
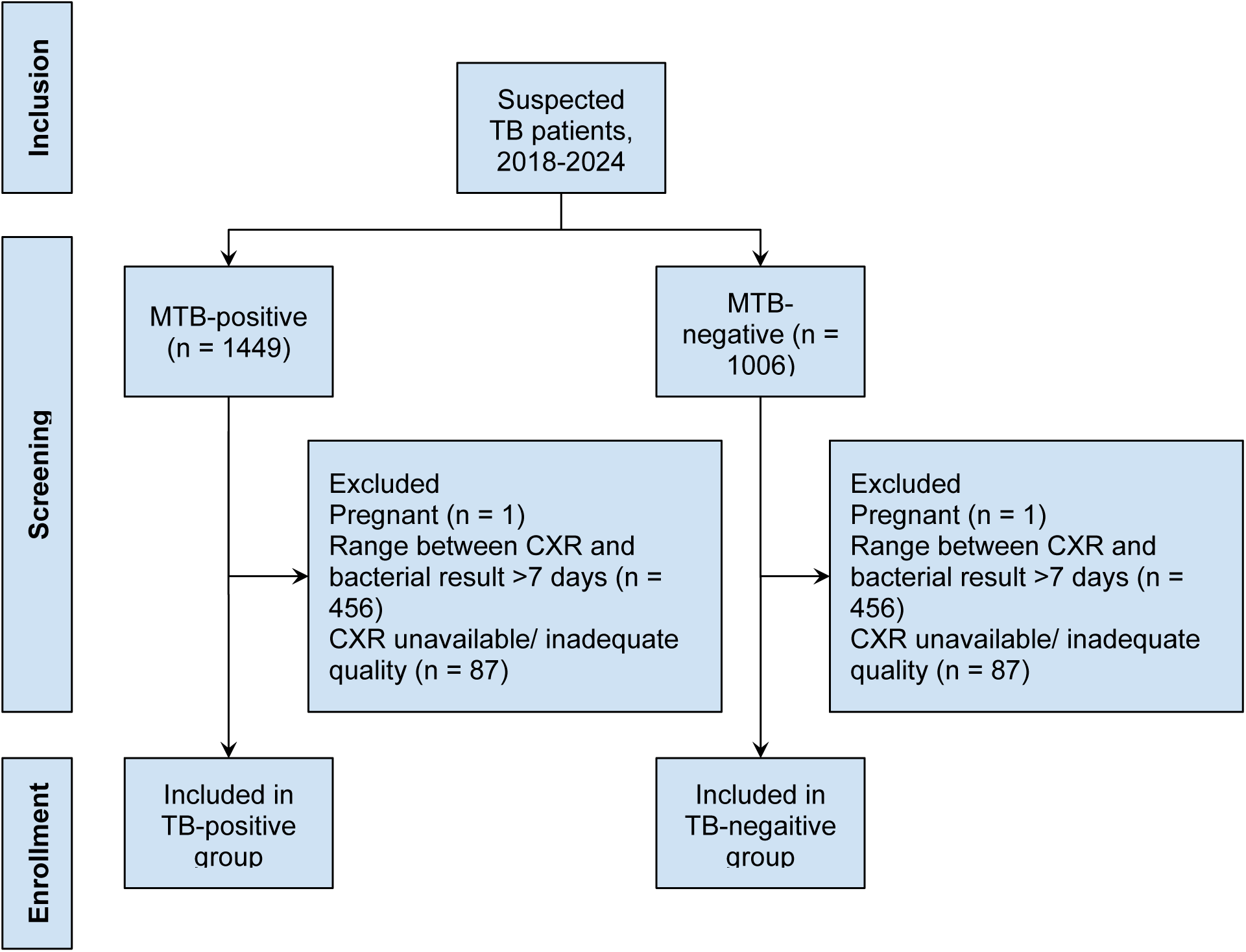
Subject flow chart during screening and enrollment phase in the study

Among the 1122 subjects, majority were male (699 subjects, 62.30%). The combined average age of subjects was 49±16 years old. As much as 40 (3.57%) subjects were human immunodeficiency virus (HIV) positive and 250 (22.28%) had a prior history of TB. Any symptom was reported in 882 subjects (78.61%), of which 703 subjects (62.66%) reported any symptom which is the part of WHO-recommended four symptoms screening (W4SS). The most common W4SS symptom was cough, that was found in 637 subjects (56.77%), followed by weight loss, reported in 271 subjects (24.15%). Other symptoms, which are not the part of W4SS, were reported by 718 subjects (63.99%), consisting of various respiratory and non-respiratory symptoms, such as shortness of breath, chest pain, malaise, decreased appetite, nausea, vomiting, and others. Table 1 further describes the subjects’ characteristics overall and in each group.

**Table 1.**
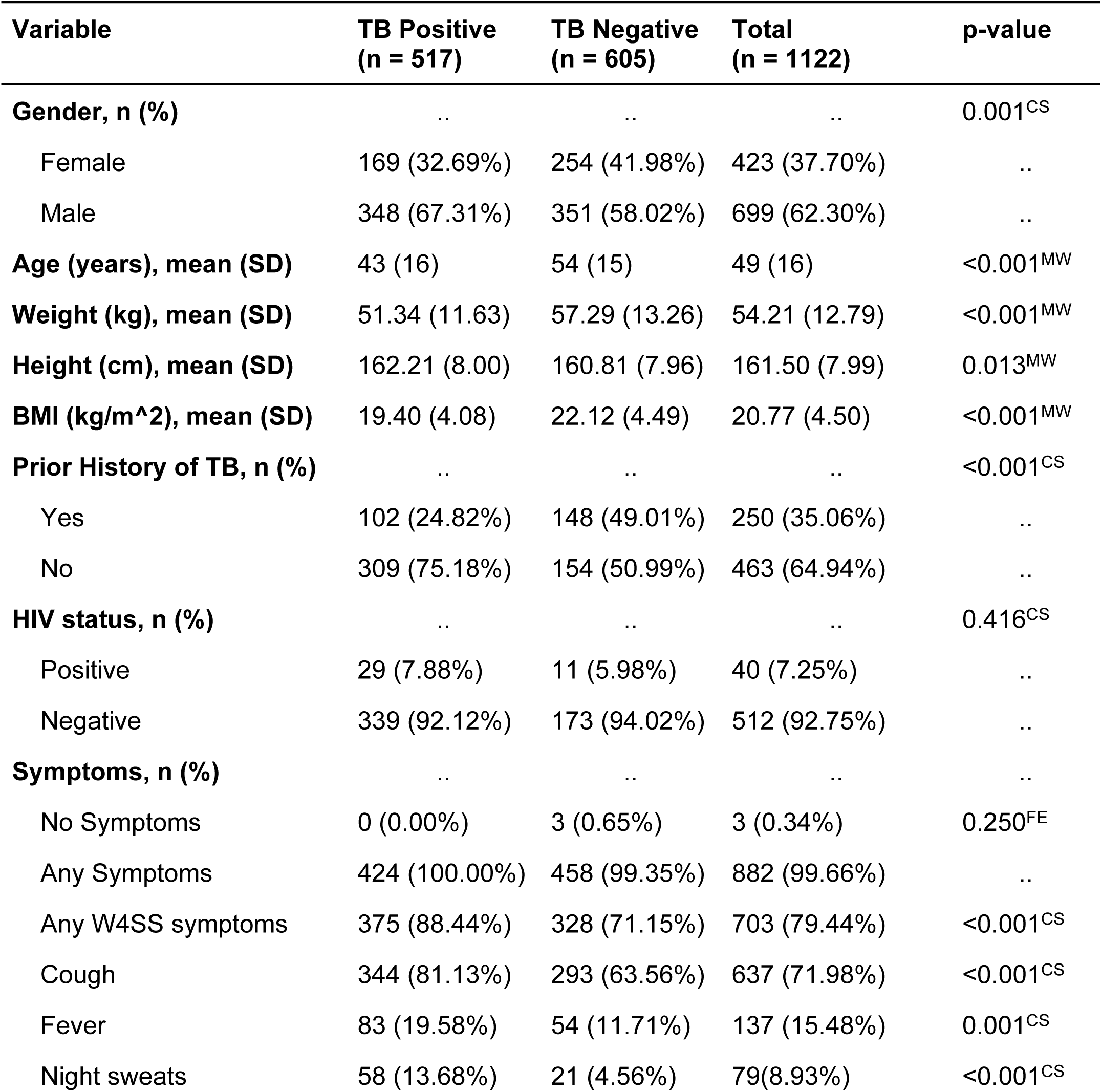
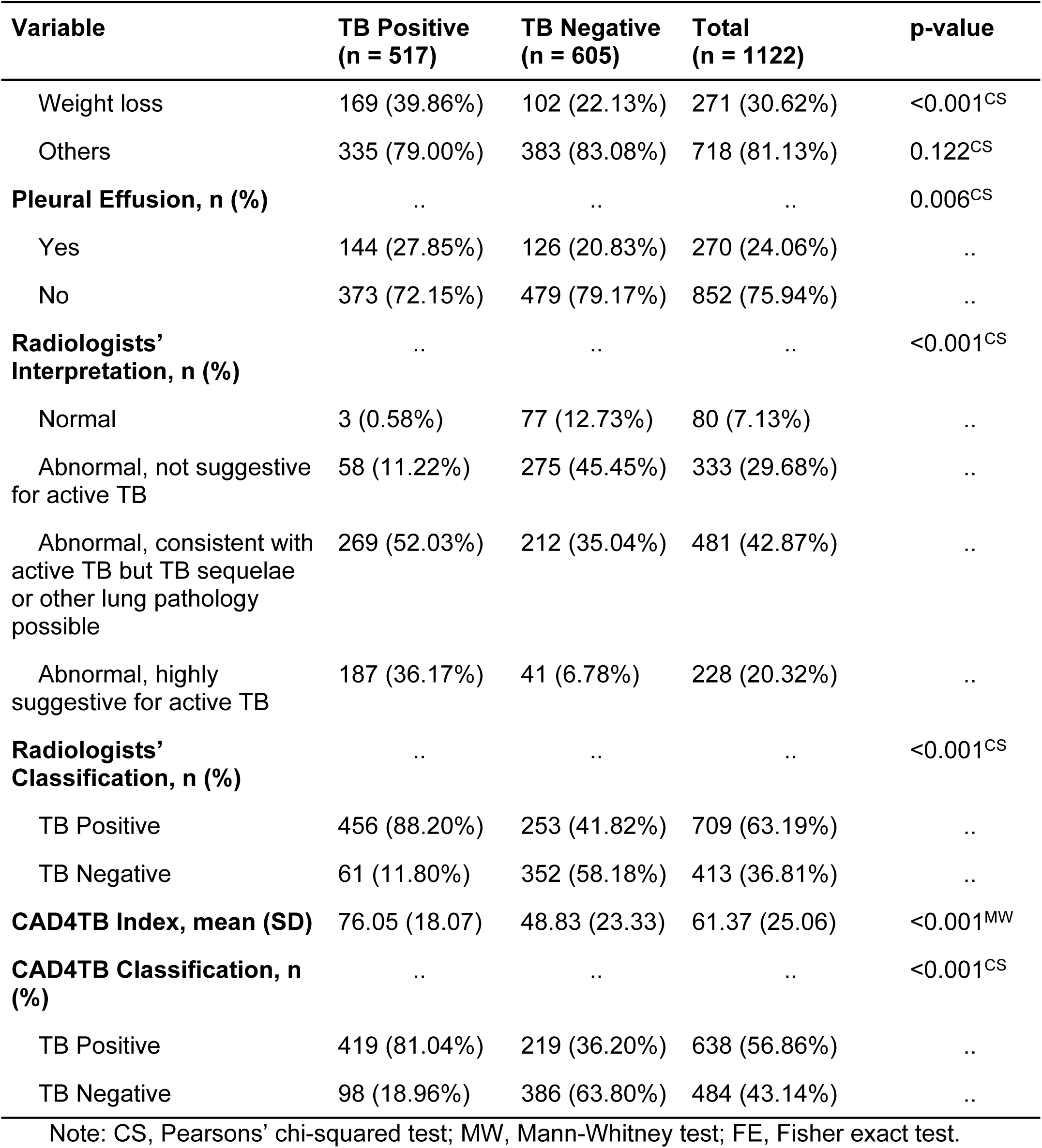
Characteristics of Participants.

### B. Diagnostic Accuracy of CAD4TB Compared with Radiologists’ in Predicting Pulmonary TB

The diagnostic accuracies of CAD4TB and radiologists’ interpretation are demonstrated through ROC curve (Figure 2) and various parameters in Table 2 below. The area under the curve (AUC) from the ROC analysis for detecting bacteriologically confirmed pulmonary TB was 0.832 (95%CI 0.808-0.856), indicating that CAD4TB has a good capability for identifying bacteriologically diagnosed pulmonary TB in this study population. Meanwhile, the AUC of radiographer interpretation scoring was 0.783 (95%CI 0.759-0.808). DeLong test of AUC paired comparison found a significant difference of AUC between CAD4TB and radiographer interpretation scoring (p-value < 0.001, 95%CI of AUC difference 0.025-0.072).

**Figure 2.**
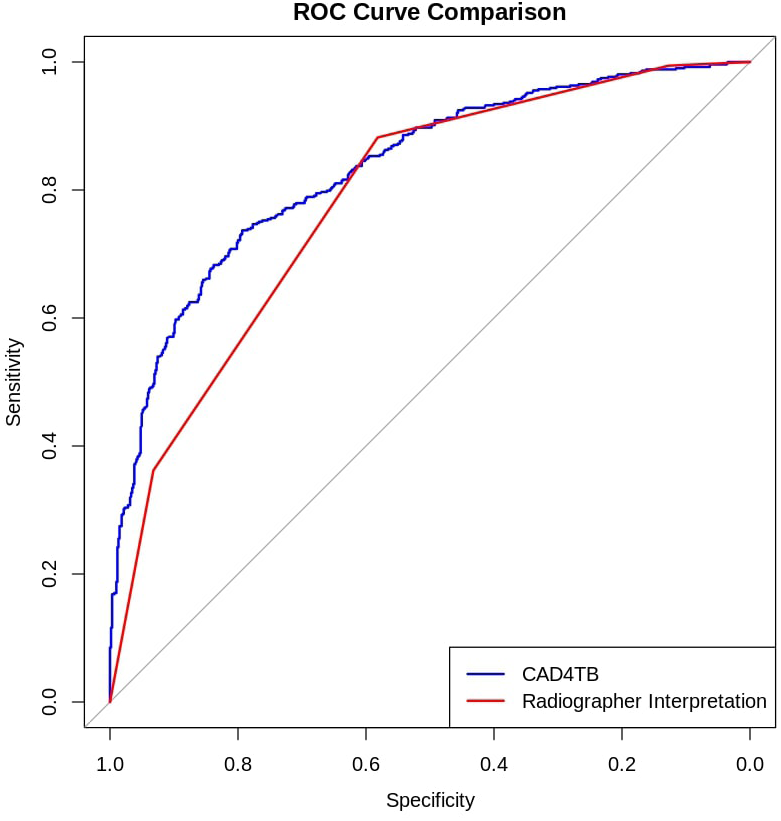
ROC curve of CAD4TB diagnostic capability and radiologists’ interpretation.

**Table 2.** Comparison of Diagnostic Parameters of CAD4TB (at Cut-off 60) and Radiologists’ Interpretation.

| Diagnostic Parameters | All Subjects |  |  |
| --- | --- | --- | --- |
|  | CAD4TB | Radiologists' Interpretation | p-value |
| Sensitivity | 81.04% (77.67%-84.42%) | 88.20% (85.42%-90.98%) | <0.001 |
| Specificity | 63.80% (59.97%-67.63%) | 58.18% (54.25%-62.11%) | <0.001 |
| PPV | 65.67% (61.99%-69.36%) | 64.32% (60.79%-67.84%) | 0.033 |
| NPV | 79.75% (76.17%-83.33%) | 85.23% (81.81%-88.65%) | <0.001 |
| PDLR | 2.24 (2.00-2.51) | 2.11 (1.91-2.33) | 0.036 |
| NDLR | 0.30 (0.25-0.36) | 0.20 (0.16-0.26) | <0.001 |

The blue line shows the sensitivity/1-specificity of CAD4TB when using different thresholds to determine TB positive result. The red line shows the sensitivity/1-specificity of radiologists’ interpretation when using different thresholds to determine TB positive result.

For CAD4TB, the analysis was conducted using the previously established cut-off of 60. Overall, sensitivity, specificity, PPV, NPV, positive and negative LR significantly differed between CAD4TB and radiologists’ interpretation, as presented in Table 2. While CAD4TB has lower sensitivity (81.04% vs 88.20%, p<0.001), it has significantly higher specificity compared to radiologists’ interpretation (63.80% vs 58.18%, p<0.001). Therefore, CAD4TB could give additional value in confirming subjects with high susceptibility of TB by radiological finding.

Based on the ROC curve analysis, we found that in the cut-off value of 53.300 the sensitivity of CAD4TB is similar with the radiologists’ interpretation, while the specificity of CAD4TB is similar with the radiologists’ interpretation in the cut-off value of 55.685. Nevertheless, the other diagnostics parameters in both cut-offs were still significantly different. In the cut-off value of 53.300, CAD4TB has lower specificity in comparison with radiologists’ interpretation (54.21% vs 58.18%, p<0.001, Table S1), while it has lower sensitivity compared to radiologists’ interpretation in the cut-off value of 55.685 (85.30% vs 88.20%, p<0.001, Table S2).

### C. Subgroup Analysis of CAD4TB

Subgroup analysis was conducted according to the existence of pleural effusion, prior history of TB, and HIV status. There was no difference of AUC and specificity between both modalities in the pleural effusion subgroup, albeit lower sensitivity of CAD4TB (p=0.281). However, in the non-pleural effusion subgroup, CAD4TB has significantly higher AUC (0.844 vs 0.794, p<0.001) and higher specificity, despite lower sensitivity (Table S3-S5, Figure S1).

In the subjects with prior TB history, while sensitivity of CAD4TB is lower, its AUC and specificity exceed radiologists’ interpretation (AUC 0.780 vs 0.681, p<0.001). Meanwhile, in the subjects without prior TB history, albeit no significant difference of specificity and AUC (p=0.119), CAD4TB has lower performance of sensitivity compared to radiologists’ interpretation (Table S6-S8, Figure S2).

Specifically in HIV-positive subjects, there was no difference of all diagnostic parameters, including AUC, between CAD4TB and radiologists’ interpretation (Table S10). On the other hand, in the HIV-negative subjects, CAD4TB has higher AUC (0.836 v 0.772, p<0.001) and better specificity (Table S9-S11, Figure S3). In conclusion, CAD4TB has comparable or better specificity against radiologists’ interpretation, especially in population without pleural effusion, with prior TB history, or with HIV-negative status.

## Discussion

### A. The Use of AI for Tuberculosis Screening

The use of artificial intelligence (AI) to assist in various tasks has become increasingly common, including the interpretation of TB screening and diagnosis using CXRs. Computer-Aided Detection for Tuberculosis (CAD4TB) is one such example. This tool works by inspecting CXRs, followed by normalization and quality assessment. CXRs that do not meet quality standards are rejected. The system then segments the lung fields and analyzes the texture. CAD4TB identifies abnormalities potentially indicative of TB using deep learning techniques on diverse CXR images from multiple countries. This analysis generates a score from 0 to 100, with 0 being normal and 100 being highly abnormal, reflecting the likelihood of TB presence. In addition to the score, the tool produces a heatmap highlighting potential abnormal areas with a color overlay on the CXR.[3]

### B. CAD4TB capability: A tale of four patients

The capability of CAD4TB in detecting TB-related lession is demonstrated in the following patient exhibiting symptoms such as cough ≥2 weeks, productive cough, bloody cough, chest pain, night sweats, and nausea. The patient underwent a mRD test, which yielded MTB-Positive and Rifampicin-Sensitive results, categorizing the patient as TB positive. The CAD4TB system assigned a score of 74.840 (Figure 3(A)), classifying this as a true positive case.

**Figure 3.**
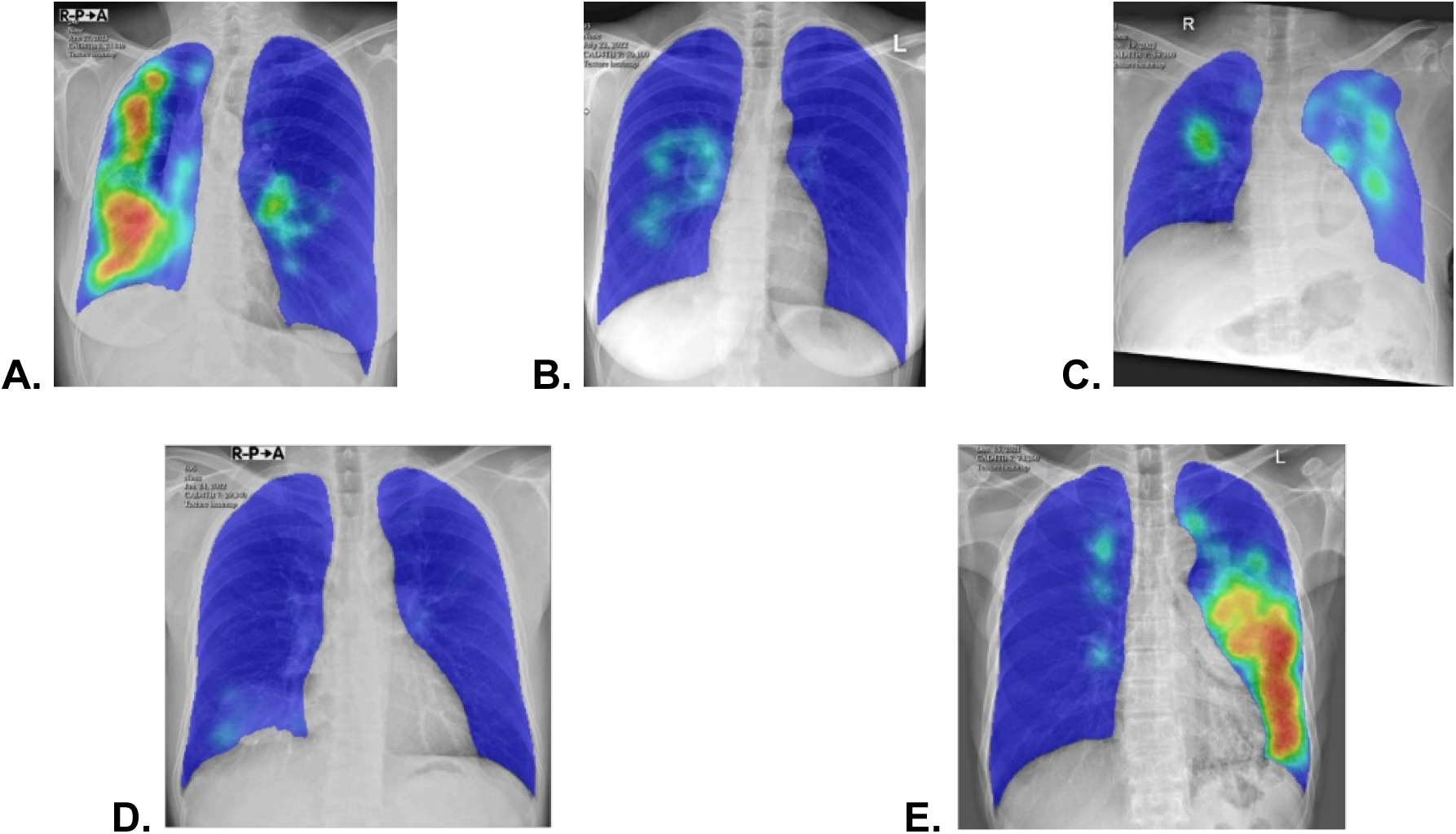
CAD4TB heatmap of CXR image in various patients: TB-positive patient (A); false-negative patient (B, C); true-negative patient, diagnosed as lung cancer (D); false-positive patient, actually diagnosed as lung cancer (E)

However, there are instances where patients suspected of having TB and who have positive bacteriological results receive a score of <60 from CAD4TB, resulting in false negative cases. For example, one of the patients had positive bacteriological results but received CAD4TB scores of 50.100 (Figure 3(B)), leading to negative diagnoses. Another example of false negative patient was a TB patient with pleural effusion (Figure 3(C))

In the group with bacteriologically negative TB, CAD4TB accurately identifies cases. For example, the patient with CXR in Figure 5(A) presented with symptoms of cough lasting ≥2 weeks, productive cough, and hemoptysis. Bacteriological assay showed negative results. CAD4TB generated an index score of 29.340 (Figure 3(D)), which is classified as a true negative, aligning with the diagnosis of lung cancer.

However, there are still cases where CAD4TB incorrectly diagnoses patients as positive for TB. For example, one of the patients presented with symptoms of shortness of breath, chest pain, cough lasting >2 weeks, productive cough, and hemoptysis. CAD4TB assessed the patient as TB positive with an index score of 74.260 (Figure 3(E)), despite the patient’s bacteriological results being negative and the actual diagnosis being a lung tumor.

### C. CAD4TB diagnostic accuracy

According to the literature, the minimum threshold for sensitivity and specificity for a screening tool is above 80%.[11] In this study, CAD4TB’s sensitivity exceeds the minimum threshold at 81.04%, indicating its efficacy in identifying true positive cases, which is crucial for a screening tool. However, its specificity of 63.80% falls below the preferred threshold. This highlights the need for confirmatory testing for patients who screen positive to ensure the accuracy of diagnosis.

Nonetheless, according to WHO recommendations, the sensitivity for screening suspected TB patients in adults using community-based triage should be 90%.[12] To achieve this level of sensitivity, the cutoff index for categorizing TB as positive in the CAD4TB interpretation needs to be set at 50.35.

The PPV and NPV were 65.67% and 79.75%, respectively. These results are within an acceptable range for a screening tool, particularly given the high prevalence of TB in Indonesia, where the priority is to avoid missing any cases. Receiver Operating Characteristic (ROC) curve analysis showed an AUC of 0.832, indicating good diagnostic accuracy, as an AUC above 0.8 is considered the minimum for a good value.[13] Another study aligned with our study and was able to distinguish between culture positive positive TB cases and non TB patients with an area under the curve of 0.84 using CAD4TB.[5] Overall, CAD4TB has good sensitivity, making it a valuable tool for TB screening, particularly in resource-limited areas due to its portability and rapid results. However, the lower specificity suggests that patients screened with this tool should undergo confirmatory testing to reduce false positives and ensure appropriate management.

### D. CAD4TB vs Radiologists’ Ability to Predict Diagnosis

This study found the significantly better diagnostic accuracy of CAD4TB compared to radiographic interpretation based on the ROC analysis. This finding reflects the ability of CAD4TB to provide similar, if not better, discrimination ability to differentiate patients with and without TB. Murphy et al,[14] with a different approach towards establishing agreement between CAD4TB and radiologists’ interpretation, also finds that CAD4TB and interpretation of radiologists were largely in agreement.

However, we also found that in the predetermined cutoff of 60, CAD4TB has inferior sensitivity, albeit with better specificity in comparison with radiologists’ interpretation. If we see from the perspective of TB screening, it can be interpreted that there are more TB cases being missed by CAD4TB-based screening compared to screening by radiologists. However, it should be noted that this study was conducted in a referral hospital in which the suspected TB patients went through the process of diagnostic confirmation. Considering this study found that CAD4TB having higher specificity than interpretation by radiologists, CAD4TB could provide additional value in identifying cases with high probability of TB in hospital settings, albeit other pulmonary conditions possibly be found in CXR imaging of hospital patients could be misinterpreted as TB could limit CAD4TB specificity.[15]

### E. Subgroup Analysis

Excluding patients with pleural effusion improved the sensitivity of both CAD4TB and radiologists, indicating that pleural effusion interferes with diagnostic accuracy. A heatmap (Figure 3(C)) illustrates a false-negative case where CAD4TB underestimated abnormalities in a TB-positive patient with massive pleural effusion (score: 59.3). This suggests that pleural effusion disrupts CAD4TB’s ability to assess lung field opacities, resulting in lower abnormality scores.

Beyond pleural effusion, other clinical conditions may influence TB-related radiographic findings. Patients with prior TB history often present with residual abnormalities, complicating diagnosis.[16] A study by Mateyo et al.[17] found that 91.83% of such patients exhibited radiographic abnormalities, regardless of TB status. Similarly, TB imaging in HIV-positive patients differs from typical cases, as common immune responses, like granuloma formation and cavitation, are often absent,[18] while atypical findings such as infiltrates are more frequent.[19]

In our study, both CAD4TB and radiologists performed worse in patients with prior TB history, consistent with previous findings.[20] CAD4TB had significantly lower sensitivity than radiologists in both subgroups, but its specificity and AUC were significantly higher in patients with prior TB history, suggesting that pre-existing abnormalities may affect radiologists more.

Interestingly, both modalities showed better diagnostic accuracy in HIV-positive patients, with higher specificity but lower sensitivity. However, the small number of HIV-positive cases in our study may explain this variation. While some studies suggest HIV-related radiographic differences impact diagnosis, others report minimal differences in CAD-based interpretations between HIV-positive and negative populations.[20,21] Further research with larger sample sizes is needed to clarify these findings.

### F. Study Strength and Limitation

A key strength of this study is its setting in a national referral hospital, allowing for the inclusion of advanced TB cases with complex radiological abnormalities. This unique population provides valuable insights into severe and atypical presentations often underrepresented in community studies. Additionally, this is the first study to compare CAD4TB analysis with radiologists’ interpretations across subgroups (pleural effusion, prior TB history, and HIV patients), highlighting CAD4TB’s strengths and weaknesses in diverse conditions.

However, some limitations should be noted. The retrospective design and reliance on secondary data from the SITB system may introduce selection bias and limit generalizability. The exclusion of certain groups, such as pregnant patients, further narrows the study’s applicability. Lastly, while CAD4TB showed promising sensitivity, its specificity remained below the optimal threshold, emphasizing the need for confirmatory testing. Future research in broader populations is needed to validate these findings.

## Conclusion

CAD4TB exhibits a promising ability to predict tuberculosis, even in hospital patients’ population with various clinical severity. With its advantages of time-effectiveness and applicability, implementation of CAD4TB in hospital settings could be considered, especially in assisting radiologists in high burden centers. Further study could be focused on developing a triage system using CAD4TB as part of TB diagnostic algorithm in hospital settings, especially considering various patient clinical condition.

## Acknowledgment

The authors would like to gratefully acknowledge the assistance of Mr. Annila Suryo Saputro from the Radiology Department of RS Persahabatan and Mr. Andiko Purnomo data officer from the tuberculosis information system (SITB) database, for their help in the data collection process. We also extend our thanks to Ardacandra Subiantoro for his contributions to data cleaning.

## Data Availability

The study data is available for sharing upon reasonable request to corresponding author. Access to the data may require institutional approval and compliance with ethical regulations.

## Supporting Information

Table S1. Comparison of Diagnostic Parameters of CAD4TB (at Cut-off 53.300) and Radiologists’ Interpretation

Table S2. Comparison of Diagnostic Parameters of CAD4TB (at Cut-off 55.685) and Radiologists’ Interpretation

Table S3. AUC of CAD4TB and radiologists’ interpretation in subjects with and without pleural effusion

Table S4. Subgroup Analysis Comparison of Diagnostic Parameters of CAD4TB (at Cut-off 60) and Radiologists’ Interpretation, Only Subjects with Pleural Effusion

Table S5. Subgroup Analysis Comparison of Diagnostic Parameters of CAD4TB (at Cut-off 60) and Radiologists’ Interpretation, Only Subjects without Pleural Effusion

Table S6. AUC of CAD4TB and radiologists’ interpretation in subjects with and without prior history of TB

Table S7. Subgroup Analysis Comparison of Diagnostic Parameters of CAD4TB (at Cut-off 60) and Radiologists’ Interpretation, Only Subjects with Prior History of TB

Table S8. Subgroup Analysis Comparison of Diagnostic Parameters of CAD4TB (at Cut-off 60) and Radiologists’ Interpretation, Only Subjects without Prior History of TB

Table S9. AUC of CAD4TB and radiologists’ interpretation in HIV-positive and HIV-negative subjects

Table S10. Subgroup Analysis Comparison of Diagnostic Parameters of CAD4TB (at Cut-off 60) and Radiologists’ Interpretation, Only HIV-Positive Subjects

Table S11. Subgroup Analysis Comparison of Diagnostic Parameters of CAD4TB (at Cut-off 60) and Radiologists’ Interpretation, Only HIV-Negative Subjects

Figure S1. ROC curve of CAD4TB and radiologists’ interpretation in subgroup with pleural effusion (A) and without pleural effusion (B)

Figure S2. ROC curve of CAD4TB and radiologists’ interpretation in subgroup with prior history of TB (A) and without prior history of TB (B)

Figure S3. ROC curve of CAD4TB and radiologists’ interpretation in HIV-positive subgroup (A) and HIV-negative subgroup (B)

